# Mother-reported EPDS-Partner (EPDS-P) Enables Indirect Screening of Paternal Perinatal Depression Independent of Maternal Factors: A Community-Based Cohort Study in Japan

**DOI:** 10.1101/2025.10.06.25337335

**Authors:** Keita Tokumitsu, Norio Sugawara, Sheehan David Fisher, Takako Keta, Junko Takeuchi, Koji Yachimori, Norio Yasui-Furukori

**Author notes:** Corresponding author address and email address: Keita Tokumitsu, Address 1: Department of Neuropsychiatry, Towada City Hospital, Towada, 034-0093, Japan, Address 2: Department of Psychiatry, Dokkyo Medical University School of Medicine, Mibu, 321-0293, Japan. Email address: Keita Tokumitsu;, Norio Sugawara, Sheehan David Fisher, Takako Keta, Junko Takeuchi, Koji Yachimori, Norio Yasui-Furukori.

## Abstract

Paternal perinatal depression negatively affects maternal mental health and child development. However, early detection remains limited, particularly in countries such as Japan, where paternal involvement in perinatal care is low. The Edinburgh Postnatal Depression Scale-Partner (EPDS-P), a maternal-rated tool for screening paternal depression, offers a feasible solution. This study longitudinally validated the diagnostic performance of the EPDS-P from the prenatal to postpartum periods and examined its independence from maternal psychological and demographic characteristics. This prospective, community-based cohort study in Towada, Japan, included couples assessed during routine visits by health nurses. Paternal depression was defined as a Center for Epidemiologic Studies Depression Scale score ≥16. Receiver operating characteristic curves were used to evaluate diagnostic accuracy, and binomial logistic regression was used to assess the influence of maternal and paternal age, parity, gestational weeks or postpartum days, and maternal Edinburgh Postnatal Depression Scale and Mother-to-Infant Bonding Scale scores. Data from 385 prenatal and 411 postpartum couples were analyzed. The EPDS-P demonstrated fair diagnostic accuracy (area under the curve: 0.783, prenatal; 0.746, postpartum). The optimal prenatal and postpartum cut-off values (3 and 4, respectively) achieved sensitivities of 70.7% and 61.5% and specificities of 75.9% and 86.8%, respectively. The EPDS-P score was the only significant predictor of paternal depression. The EPDS-P is a valid and practical tool for indirectly screening for paternal perinatal depression, independent of maternal influences. Its integration into routine perinatal care may help identify at-risk fathers in settings with limited paternal engagement and contribute to more inclusive, family-centered mental health support.

**Highlights:**

- The EPDS-P enables maternal indirect screening of paternal perinatal depression.
- A cohort of 385 prenatal and 411 postpartum Japanese couples were surveyed.
- The EPDS-P accuracy was fair, with AUCs of 0.783 (prenatal) and 0.746 (postpartum).
- The optimal EPDS-P cutoffs were 3 (prenatal) and 4 (postpartum).
- EPDS-P accuracy was independent of maternal mood or bonding measures.

## Introduction

Perinatal depression refers to depressive symptoms that emerge during pregnancy or within the first postpartum year and is a serious mental health concern due to its detrimental impact on both maternal well-being and child development (Stuart-Parrigon and Stuart, 2014). A range of biological, psychological, and social factors are involved in the onset of maternal perinatal depression, and family support has consistently been reported as a major protective factor against poor maternal mental health (O’Hara and Swain, 1996). Consequently, men are often expected to play a supportive role for their partners and children throughout this period (Mezulis et al., 2004).

In recent years, increasing attention has been paid to the psychological difficulties that men may encounter during the perinatal period. Research has shown that a significant number of fathers exhibit depressive symptoms, with a global meta-analysis reporting a paternal perinatal depression prevalence of approximately 8.4% (Cameron et al., 2016; Paulson and Bazemore, 2010). This prevalence rate underscores the fact that paternal depression is a significant global public health issue that is not confined to a single culture or region. Our previous study on the Japanese population yielded a comparable prevalence of approximately 10% and found that the rate of paternal postpartum depression was similar to that observed in women (Tokumitsu et al., 2020). Accumulating evidence suggests a bidirectional relationship between maternal and paternal perinatal depression, both of which are associated with a higher likelihood of child maltreatment. These findings emphasize the importance of father-targeted interventions to enhance maternal psychological stability and support optimal child development.

While maternal screening is routinely implemented during prenatal and postnatal checkups in many countries, men often have limited opportunities to attend these visits, making the early identification of at-risk fathers challenging. This challenge is exacerbated by the fact that healthcare professionals’ awareness that men can suffer from perinatal depression continues to lag (Cameron et al., 2017). Although internet-based screening and support tools offer scalable, low-burden options for engaging fathers, such approaches remain underutilized in many regions, including Japan (Hussain-Shamsy et al., 2020). These challenges highlight the urgent need for pragmatic, globally adaptable strategies to promote paternal mental health during the perinatal period.

In Japan, self-reported screening instruments, such as the Kessler Psychological Distress Scale (Kessler et al., 2002; Sakurai et al., 2011) and the Patient Health Questionnaire-9 (Spitzer et al., 1999), have shown limited validity when used by female partners as indirect measures to evaluate the mental health status of fathers, highlighting the need for more appropriate observer-rated instruments (Konishi et al., 2016). The Edinburgh Postnatal Depression Scale (EPDS), a 10-item self-reported questionnaire developed by Cox et al. (Cox et al., 1987), has been adopted worldwide to assess maternal perinatal depression, and its Japanese version has shown strong psychometric properties (Okano et al., 1996). Based on this instrument, the EPDS-Partner (EPDS-P) was developed as a screening tool based on maternal evaluations to detect depressive symptoms in fathers (Fisher et al., 2012). While the EPDS-P has demonstrated promising validity in earlier studies, these investigations were limited by small sample sizes and were confined to the postpartum period. One notable advantage of the EPDS-P is that it can indirectly assess paternal depression via maternal reports, even when fathers do not engage in support programs or during times of restricted healthcare access, such as during the COVID-19 pandemic. However, the tool’s reliability and applicability in large-scale studies and non-Western populations, including those in Japan, have not yet been thoroughly evaluated. Furthermore, no prior research has validated the EPDS-P for longitudinal use spanning both the prenatal and postnatal stages.

Additionally, although self-reported screening tools, such as the paternal EPDS, have been used to directly assess depressive symptoms in fathers (Nishimura and Ohashi, 2010), there is a lack of evidence comparing the screening accuracy of the EPDS-P and paternal EPDS within the same cohort. The paternal EPDS, as a self-rated instrument, may be influenced by cultural and gendered expectations that discourage men from expressing emotional vulnerability, potentially leading to the underreporting of symptoms. Conversely, indirect maternal raters may have underestimated paternal psychological distress. Understanding the extent of agreement and discrepancy between partner-rated and self-rated assessments is essential for designing effective, family-centered screening systems.

To address these gaps, we conducted a longitudinal study in Japan using a community-based cohort to evaluate the diagnostic accuracy and optimal cut-off scores of both the Japanese version of the EPDS-P and the paternal EPDS for identifying paternal perinatal depression. By assessing these tools at the prenatal and postnatal stages, we aimed to better understand the reliability and utility of these indirect and direct assessment methods and help develop inclusive, scalable mental health support strategies for families worldwide, especially by establishing a practical, indirect screening method applicable in diverse healthcare settings where direct paternal engagement is limited.

## Methods

### Study Design and Participants

This prospective observational study was conducted in Towada City, Aomori Prefecture, Japan. The participants were recruited during routine perinatal home visits conducted by public health nurses and midwives affiliated with the Towada City Community Health Center (Tokumitsu et al., 2023). These professionals had extensive experience in maternal and child healthcare and had completed prior training in research ethics. This naturalistic cohort included all individuals residing in the community who were in the prenatal or postpartum period during the study period. Participants were not randomized or allocated to any intervention group. All eligible individuals received identical self-administered questionnaires and psychoeducational information upon enrollment, regardless of the timing. The study documents and questionnaires were prepared in Japanese. Individuals unable to comprehend Japanese were excluded from the study. Additionally, participants under the age of 18 were excluded because of their legal inability to provide informed consent. All participants received a full explanation of the study procedure and provided written informed consent.

### Outcome Measures

The primary outcome was the screening accuracies of the Japanese version of the EPDS-P and the paternal EPDS for detecting paternal perinatal depression. Depression was defined based on the Center for Epidemiologic Studies Depression Scale (CES-D), with scores ≥16 indicating depression. Receiver operating characteristic (ROC) curves were generated, and the area under the curve (AUC) values were calculated to assess the discriminatory capacity of the EPDS-P and paternal EPDS. The optimal cut-off score was determined based on the Youden Index by identifying the point at which the sum of sensitivity and specificity (sensitivity + specificity – 1) was maximized. The sensitivity and specificity of the cut-off values were then reported. All statistical analyses were performed using SPSS Statistics version 28 (IBM Corp., Armonk, NY, USA) for Microsoft Windows and EZR (Saitama Medical Center, Jichi Medical University, Japan), a graphical interface for R version 3.5.2. EZR is a modified version of the R commanders tailored for medical statistics (Kanda, 2013). P-values of <0.05 were considered statistically significant.

### Measures

EPDS-P (Fisher et al., 2012)

The EPDS-P was originally developed by Fisher et al. (2012) to evaluate paternal postpartum depression based on maternal reporting. Although its diagnostic accuracy was reported to have 89% sensitivity and 59% specificity at a cut-off of 4/5 in English, no validated Japanese version exists. With Dr. Fisher’s permission and collaboration, the EPDS-P was translated into Japanese and back-translated by three native speakers of Japanese and English to confirm its cross-linguistic validity. Figure S1 shows the Japanese version of the EPDS-P. Its reliability was evaluated at two time points: during pregnancy and the postpartum period.

### CES-D (Radloff, 1977)

The CES-D is a 20-item self-report questionnaire developed by the U.S. National Institute of Mental Health to screen for depressive symptoms in the general population. This study used a validated Japanese version of the CES-D (Shima et al., 1985). Male participants who completed the CES-D and scored ≥16 were classified with depression.

EPDS (Cox et al., 1987)

The EPDS is a 10-item self-reported screening tool for postpartum depression developed by British psychiatrist John Cox. The Japanese version has been validated and widely used since 1996 (Okano et al., 1996). Since previous studies have suggested interrelated depressive symptoms between mothers and fathers during the perinatal period, maternal EPDS scores were included as covariates to assess their potential influence on EPDS-P performance.

Mother-to-Infant Bonding Scale (MIBS) (Taylor et al., 2005)

The MIBS is a 10-item self-reported scale used to assess maternal bonding with infants, with higher scores indicating weaker bonding. The Japanese version has been validated and widely used in Japan (Yoshida et al., 2012). Although its influence on EPDS-P accuracy has not yet been investigated, maternal MIBS scores were included as covariates to control for possible biases.

Figure 1 illustrates the direction and type of assessment for each scale, including whether it is a self- or partner-reported assessment.

**Figure 1.**
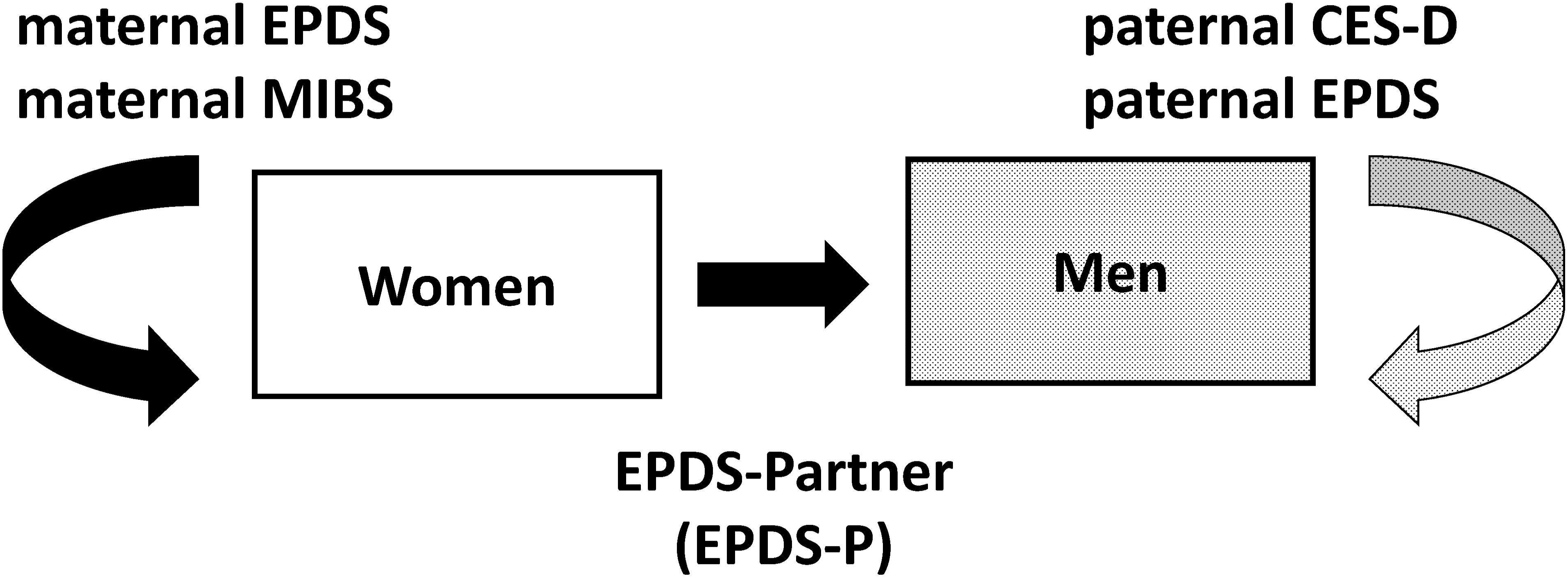
Schematic overview of the assessment procedures illustrating the different assessment modalities used in this study, including self-reported and partner-reported scales. Women completed self-reported scales to assess their psychological state (maternal EPDS and MIBS) and a partner-reported scale to assess their partners’ depressive symptoms (EPDS-P). The men completed self-reported scales to assess depressive symptoms (paternal CES-D and paternal EPDS scores). Abbreviations: CES-D: Center for Epidemiologic Studies Depression Scale; EPDS: Edinburgh Postnatal Depression Scale; EPDS-P: Edinburgh Postnatal Depression Scale-Partner version; MIBS: Mother-to-Infant Bonding Scale.

### Covariates and Sensitivity Analyses

A binomial logistic regression analysis was conducted as a sensitivity analysis to assess the robustness of EPDS-P using paternal depression (male participants with CES-D scores ≥16) as the dependent variable. The independent variables included the EPDS-P score, maternal and paternal age, parity, gestational weeks or postpartum days, and the maternal EPDS and MIBS scores. This analysis was conducted to determine whether each variable significantly predicted paternal depression.

### Ethical Approvals

This study was conducted following the Declaration of Helsinki and the Ethical Guidelines for Medical and Health Research Involving Human Subjects in Japan. The study protocol was reviewed and approved by the Ethics Committee of the Dokkyo Medical University School of Medicine (Approval No. 2021-015) (Tokumitsu et al., 2023). All participants provided written informed consent after receiving a full explanation of the study.

Administrative approval was obtained for data access. The data contained potentially identifiable or sensitive information; therefore, data sharing was restricted by the Ethics Committee. The use of this database was approved by our institutional review board. Please contact the corresponding author for requests to access the data.

## Results

This study was conducted in Towada, Aomori, Japan, from October 1, 2021, to March 31, 2025. Of the eligible participants, 540 couples had the opportunity to participate during the prenatal period, 578 during the postpartum period, and 385 couples were eligible for both periods. In total, 402 and 425 couples provided informed consent during the prenatal and postpartum periods. Valid respondents were defined as couples who provided complete data on age, sex, and both the EPDS-P and CES-D assessments; 385 couples in the prenatal period (valid response rate: 71.3%) and 411 couples in the postpartum period (valid response rate: 71.1%) were included in the analysis. A total of 267 couples provided valid responses at both time points and were included in the longitudinal analysis. Figure 2 presents a flowchart of the participant selection process.

**Figure 2.**
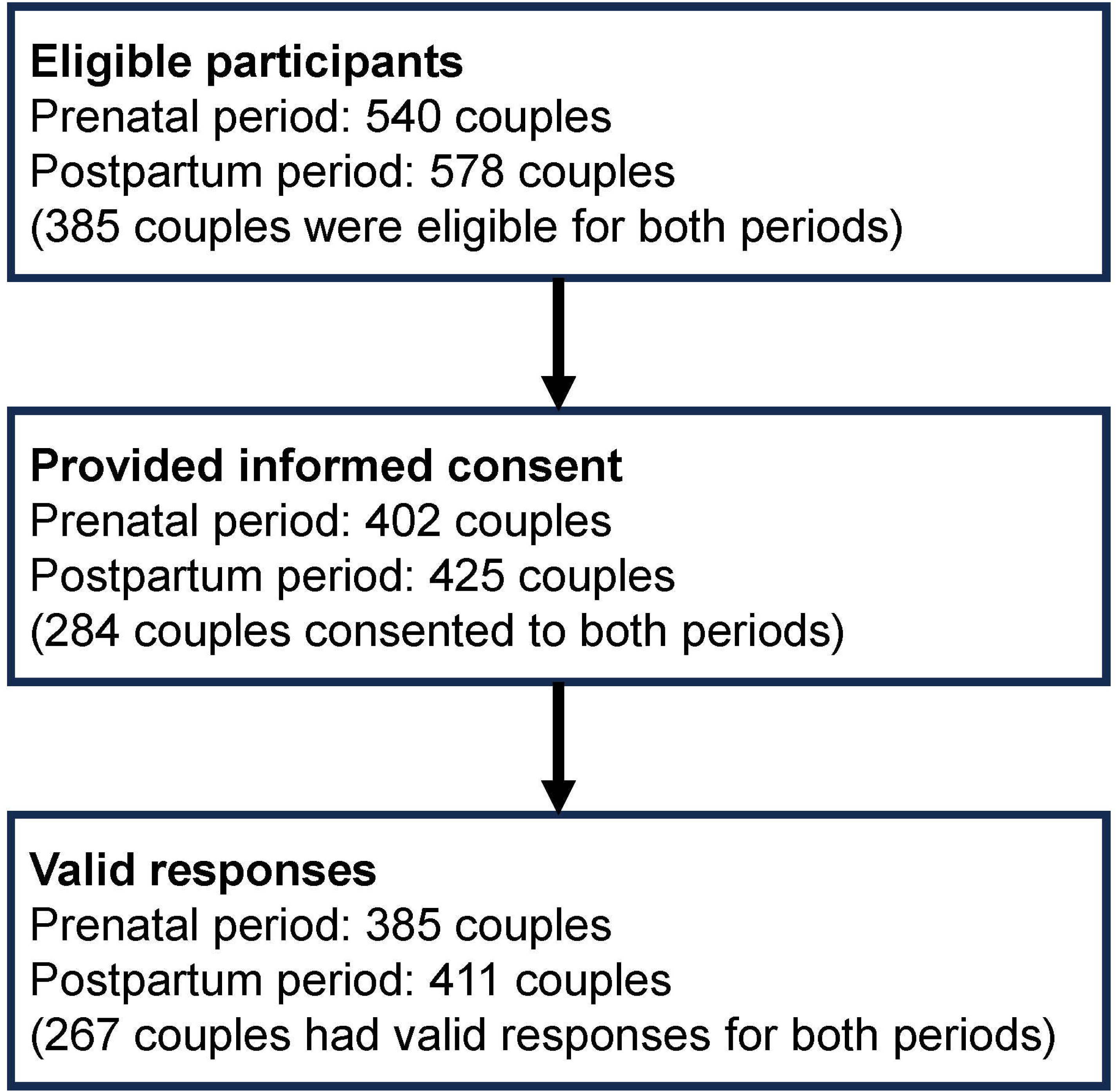
Participant flow diagram showing the number of couples at each stage of the study in the prenatal and postpartum periods. The final analysis cohort (“Valid responses”) is defined as couples who provided complete data on age, sex, and both the EPDS-P and CES-D scales. The valid response rates are 71.3% (385 of 540 eligible couples) during the prenatal period and 71.1% (411 of 578 eligible couples) during the postpartum period. Abbreviations: CES-D: Center for Epidemiologic Studies Depression Scale; EPDS-P: Edinburgh Postnatal Depression Scale-Partner.

The mean ages of the male and female participants were 33.8 and 31.3 years during the prenatal period and 34.1 and 31.5 years during the postpartum period, respectively. The proportion of first-time parents was 42.6% (164/385) among the prenatal participants and 42.3% (174/411) among the postpartum participants. The prevalence of paternal depression, assessed using the CES-D completed by the fathers (cut-off score: ≥16), was 10.6% (41/385) during the prenatal period and 6.3% (26/411) during the postpartum period. The prevalence of maternal depression, assessed using the EPDS completed by the mothers (cut-off score: ≥9), was 7.0% (27/384) during the prenatal period and 5.4% (22/408) during the postpartum period. The prevalence of depression did not differ between men and women in either period (chi-square test, p >0.05). Among the 267 couples who provided longitudinal data at both the prenatal and postpartum time points, 11 out of 14 men (78.6%) who exhibited postpartum depression were already depressed during the prenatal period. Table 1 presents the sociodemographic and psychological characteristics of the participants.

**Table 1.**
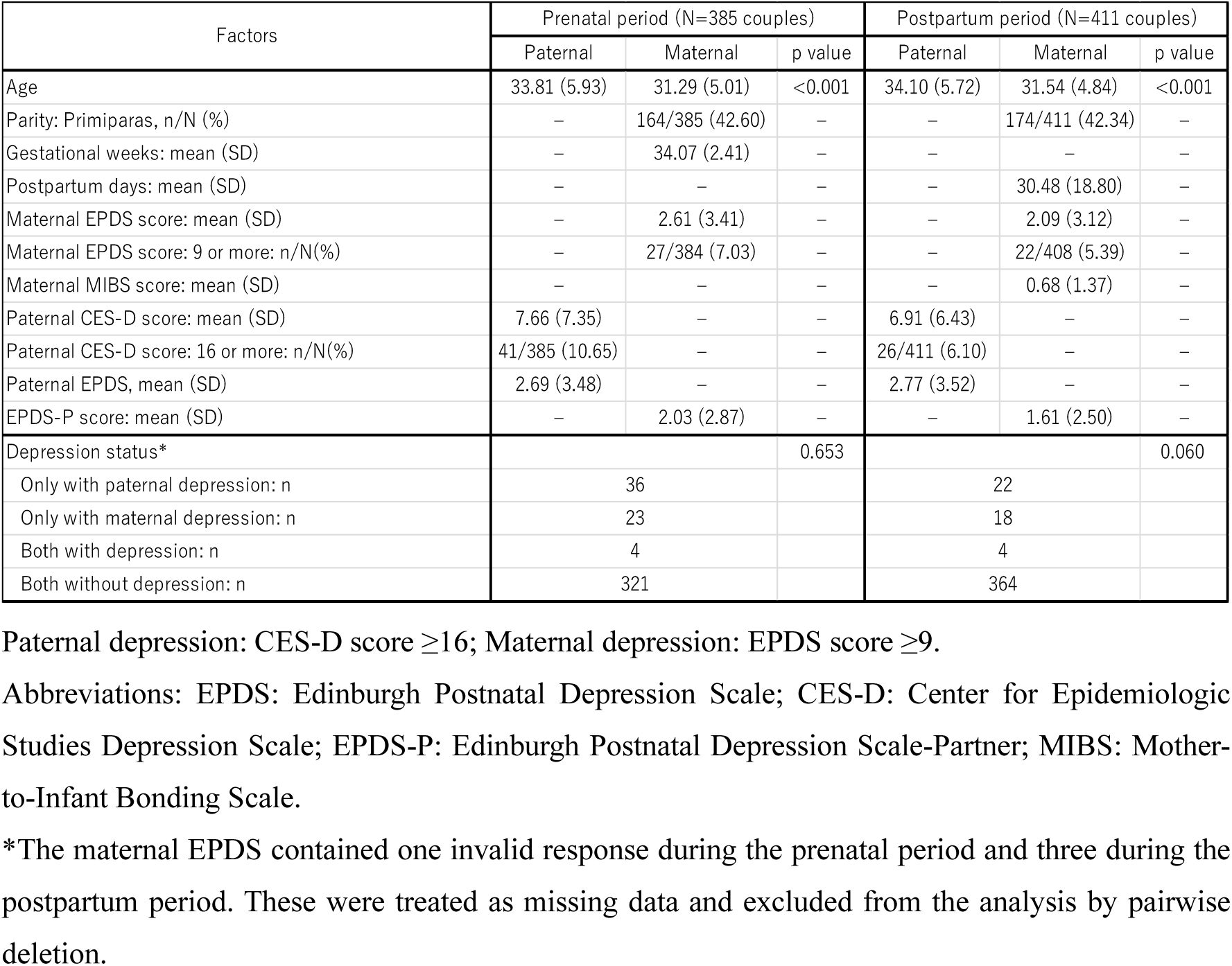
Sociodemographic and psychological characteristics of paternal and maternal participants in the prenatal and postpartum periods.

| Factors | Prenatal period (N=385 couples) |  |  | Postpartum period (N=411 couples) |  |  |
| --- | --- | --- | --- | --- | --- | --- |
|  | Paternal | Maternal | p value | Paternal | Maternal | p value |
| Age | 33.81 (5.93) | 31.29 (5.01) | <0.001 | 34.10 (5.72) | 31.54 (4.84) | <0.001 |
| Parity: Primiparas, n/N (%) | – | 164/385 (42.60) | – | – | 174/411 (42.34) | – |
| Gestational weeks: mean (SD) | – | 34.07 (2.41) | – | – | – | – |
| Postpartum days: mean (SD) | – | – | – | – | 30.48 (18.80) | – |
| Maternal EPDS score: mean (SD) | – | 2.61 (3.41) | – | – | 2.09 (3.12) | – |
| Maternal EPDS score: 9 or more: n/N(%) | – | 27/384 (7.03) | – | – | 22/408 (5.39) | – |
| Maternal MIBS score: mean (SD) | – | – | – | – | 0.68 (1.37) | – |
| Paternal CES-D score: mean (SD) | 7.66 (7.35) | – | – | 6.91 (6.43) | – | – |
| Paternal CES-D score: 16 or more: n/N(%) | 41/385 (10.65) | – | – | 26/411 (6.10) | – | – |
| Paternal EPDS, mean (SD) | 2.69 (3.48) | – | – | 2.77 (3.52) | – | – |
| EPDS-P score: mean (SD) | – | 2.03 (2.87) | – | – | 1.61 (2.50) | – |
| Depression status* |  |  | 0.653 |  |  | 0.060 |
| Only with paternal depression: n | 36 |  |  | 22 |  |  |
| Only with maternal depression: n | 23 |  |  | 18 |  |  |
| Both with depression: n | 4 |  |  | 4 |  |  |
| Both without depression: n | 321 |  |  | 364 |  |  |

Paternal depression: CES-D score $\geq 16$ ; Maternal depression: EPDS score $\geq 9$ .
Abbreviations: EPDS: Edinburgh Postnatal Depression Scale; CES-D: Center for Epidemiologic Studies Depression Scale; EPDS-P: Edinburgh Postnatal Depression Scale-Partner; MIBS: Mother-to-Infant Bonding Scale.
\*The maternal EPDS contained one invalid response during the prenatal period and three during the postpartum period. These were treated as missing data and excluded from the analysis by pairwise deletion.

The Japanese version of the EPDS-P demonstrated acceptable internal consistency, with Cronbach’s alpha coefficients of 0.767 and 0.749 for the prenatal and postpartum periods, respectively. At the optimal cut-off scores of 3 for the prenatal period and 4 for the postpartum period, the sensitivity and specificity were 70.7% and 75.9% during the prenatal period and 61.5% and 86.8% during the postpartum period, respectively. Figure 3 presents the corresponding ROC curves, and Table 2 provides the sensitivity and specificity values for various EPDS-P cut-off scores for detecting paternal perinatal depression. The scale also showed moderate but statistically significant discriminatory ability, with AUC values of 0.783 (95% confidence interval [CI]: 0.700–0.866, n = 385) during the prenatal period and 0.746 (95% CI: 0.629–0.863, n = 411) during the postpartum period. Both AUCs fell within the “fair” range (0.70–0.79), and their confidence intervals exceeded the null threshold of 0.5, indicating good discriminatory performance at both time points (Fischer et al., 2003). AUC interpretations were based on the criteria of Fischer et al. (Fischer et al., 2003). Figure 4 presents the scatter plots of the total EPDS-P and CES-D scores during the prenatal and postpartum periods. Spearman’s rank correlation revealed a moderate and significant association during the prenatal period (ρ = 0.410, p <0.001) and a weaker but still significant association during the postpartum period (ρ = 0.246, p <0.001). The correlation coefficients are presented in temporal sequence. Although our protocol initially estimated the required sample size of 554 couples per time point based on the prevalence estimation (Tokumitsu et al., 2023), the present analysis focused on evaluating the diagnostic accuracy of the EPDS-P. In this context, sample sizes exceeding 300 are considered sufficient for reliable AUC estimation, as suggested by Hajian-Tilaki (Hajian-Tilaki, 2014). Therefore, the sample sizes used in this study were deemed adequate for validating the EPDS-P.

**Figure 3.**
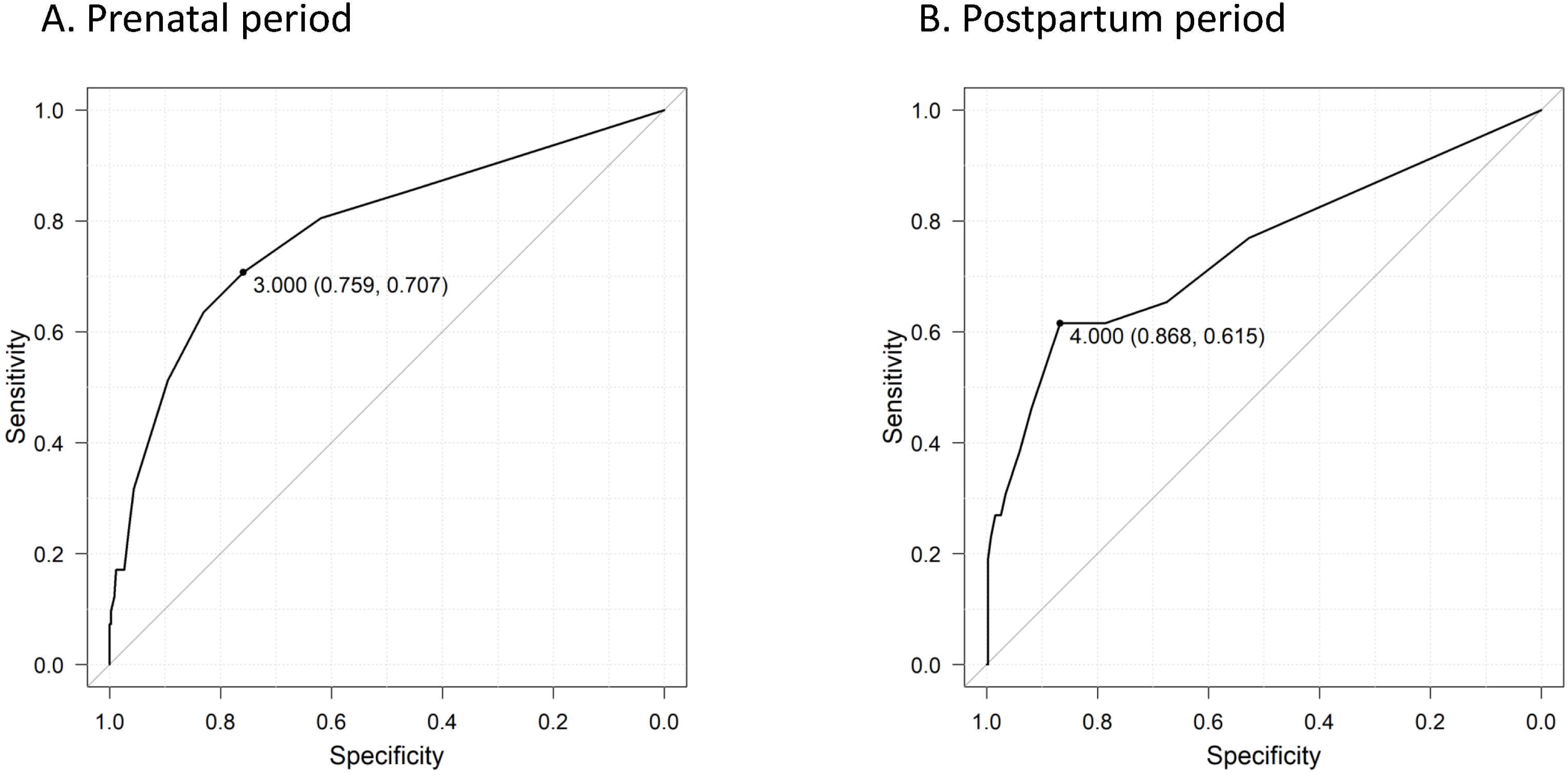
ROC curves of the EPDS-P for detecting paternal perinatal depression (CES-D ≥16) during the prenatal and postpartum periods. The curves illustrate the diagnostic performance of the EPDS-P for (A) the prenatal period (AUC = 0.783; 95% CI, 0.700–0.866) and (B) the postpartum period (AUC = 0.746; 95% CI, 0.629–0.863). The marked points indicate the optimal cut-off scores determined by the Youden Index, with coordinates presented as (Specificity, Sensitivity). The diagonal grey line represents the no-discrimination line. Abbreviations: AUC, area under the curve; CI, confidence interval; CES-D, Center for Epidemiologic Studies Depression Scale; EPDS-P, Edinburgh Postnatal Depression Scale–Partner; ROC, receiver operating characteristic

**Figure 4.**
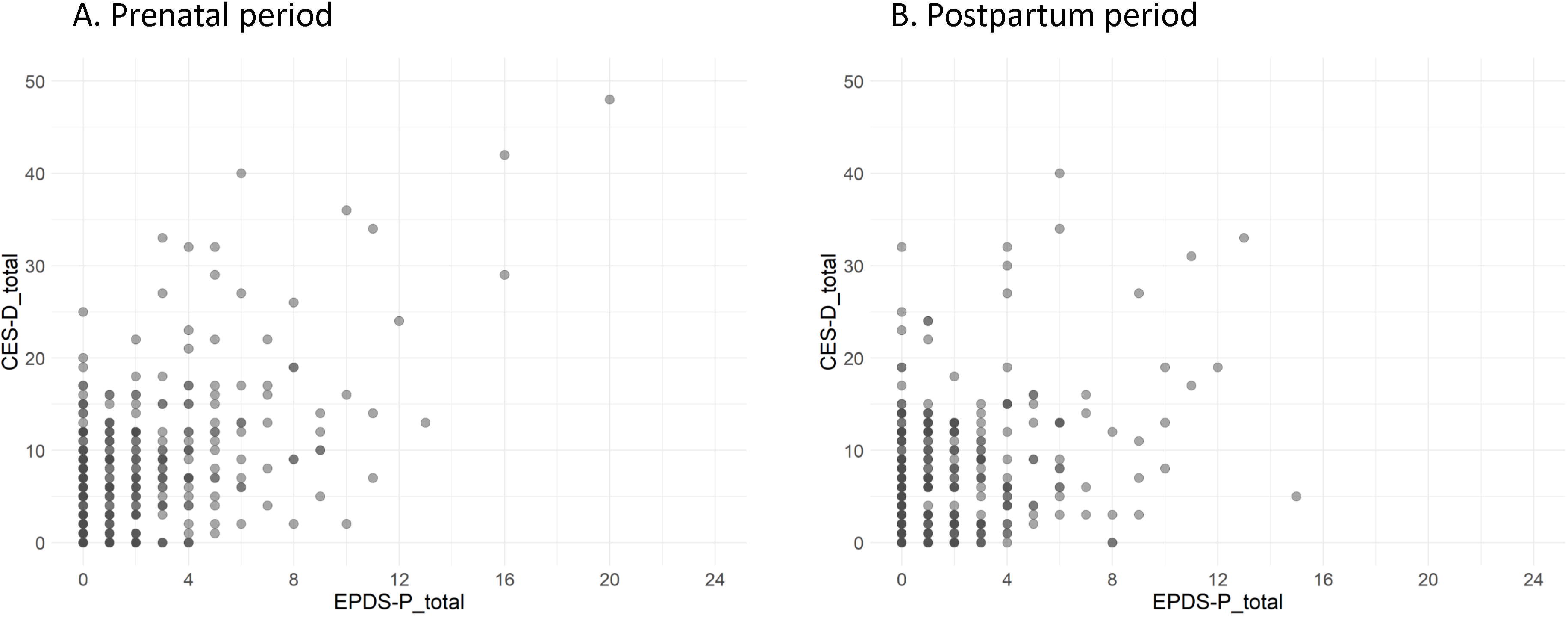
Scatter plots of the EPDS-P scores versus paternal CES-D scores during the prenatal and postpartum periods illustrating the relationship between the EPDS-P (completed by mothers) and CES-D (completed by fathers) for the (A) prenatal and (B) the postpartum periods. Each point represents an individual participating couple. Spearman’s rank correlation reveals a moderate, significant association during the prenatal period (ρ = 0.410, p <0.001) and a weaker but significant association during the postpartum period (ρ = 0.246, p <0.001). CES-D: Center for Epidemiologic Studies Depression Scale; EPDS-P: Edinburgh Postnatal Depression Scale-Partner. Abbreviations: CES-D: Center for Epidemiologic Studies Depression Scale; EPDS-P: Edinburgh Postnatal Depression Scale–Partner.

**Table 2.** Sensitivity and specificity of the EPDS-P at various cutoff scores for detecting paternal perinatal depression (CES-D ≥16) during the prenatal and postpartum periods.

| EPDS-P Cutoff Score | Prenatal period |  | Postpartum period |  |
| --- | --- | --- | --- | --- |
|  | Sensitivity | Specificity | Sensitivity | Specificity |
| $\geq 0$ | 100.0% | 0.0% | 100.0% | 0.0% |
| $\geq 1$ | 85.4% | 46.2% | 76.9% | 52.7% |
| $\geq 2$ | 80.5% | 61.9% | 65.4% | 67.5% |
| $\geq 3$ | 70.7% | 75.9% | 61.5% | 78.7% |
| $\geq 4$ | 63.4% | 83.1% | 61.5% | 86.8% |
| $\geq 5$ | 51.2% | 89.5% | 46.2% | 91.9% |
| $\geq 6$ | 39.0% | 93.3% | 38.5% | 94.0% |
| $\geq 7$ | 31.7% | 95.6% | 30.8% | 96.6% |
| $\geq 8$ | 24.4% | 96.5% | 26.9% | 97.4% |
| $\geq 9$ | 17.1% | 97.4% | 26.9% | 98.4% |
| $\geq 10$ | 17.1% | 98.8% | 23.1% | 99.2% |
| $\geq 11$ | 12.2% | 99.1% | 19.2% | 99.7% |
| $\geq 12$ | 9.8% | 99.7% | 11.5% | 99.7% |
| $\geq 13$ | 7.3% | 99.7% | 7.7% | 99.7% |
| $\geq 14$ | – | – | 3.8% | 99.7% |
| $\geq 15$ | – | – | 0.0% | 99.7% |
| $\geq 16$ | 7.3% | 100.0% | – | – |
| $\geq 20$ | 2.4% | 100.0% | – | – |

“–” Values not calculable due to the absence of participants with EPDS-P scores at those thresholds during the prenatal period.
The optimal cut-off scores were $\geq 3$ for the prenatal period and $\geq 4$ for the postpartum period, as determined by the Youden index.
Abbreviations: CES-D: Center for Epidemiologic Studies Depression Scale; EPDS-P: Edinburgh Postnatal Depression Scale-Partner.

For the sensitivity analysis, a binomial logistic regression analysis was performed with paternal depression as the dependent variable and the EPDS-P score as the independent variable. The covariates included maternal and paternal age, parity, gestational weeks or postpartum days, and the maternal EPDS and MIBS scores. The EPDS-P scores were significantly associated with paternal depression during the prenatal and postpartum periods. However, none of the covariates showed statistically significant associations. Table 3 presents these results.

**Table 3.** Adjusted ORs and 95% CIs from logistic regression analyses for paternal depression in the prenatal and postpartum periods.

| Factors | Model 1 (Prenatal) |  | Model 2 (Postpartum) |  |
| --- | --- | --- | --- | --- |
|  | OR (95% CI) | P value | OR (95% CI) | P value |
| Maternal age | 1.009 (0.917–1.110) | 0.851 | 0.944 (0.820–1.086) | 0.417 |
| Paternal age | 0.975 (0.902–1.054) | 0.517 | 0.976 (0.874–1.089) | 0.663 |
| Parity: Multiparas (Ref = Primiparas) | 0.629 (0.289–1.370) | 0.243 | 1.123 (0.436–2.891) | 0.811 |
| Gestational weeks | 0.959 (0.823–1.117) | 0.588 | – | – |
| Postpartum days | – | – | 1.013 (0.995–1.032) | 0.161 |
| Maternal EPDS score | 0.959 (0.857–1.073) | 0.461 | 1.039 (0.903–1.195) | 0.595 |
| Maternal MIBS score | – | – | 1.241 (0.947–1.627) | 0.117 |
| EPDS-P score | 1.415 (1.261–1.589) | <0.001 | 1.385 (1.220–1.571) | <0.001 |
| Constant | 0.440 | 0.776 | 0.164 | 0.215 |

Paternal depression: CES-D $\geq 16$ .
One case in the prenatal model and three in the postpartum model were excluded because of missing maternal EPDS data, resulting in 384 and 408 couples, respectively.
Abbreviations: EPDS: Edinburgh Postnatal Depression Scale; CES-D: Center for Epidemiologic Studies Depression Scale; CI: Confidence interval; EPDS-P: Edinburgh Postnatal Depression Scale-Partner; MIBS: Mother-to-Infant Bonding Scale; OR: Odds ratio.

As an additional analysis, we examined the diagnostic accuracy and optimal cut-off score of the Japanese version of the paternal EPDS for detecting paternal perinatal depression in Japanese men. Among the participants who provided informed consent, 387 men in the prenatal period and 412 men in the postnatal period completed both the paternal EPDS and CES-D without missing data. The paternal EPDS demonstrated acceptable internal consistency, with Cronbach’s alpha coefficients of 0.817 and 0.822 in the prenatal and postpartum periods, respectively. At the optimal cut-off score of 4 for both time points, the sensitivity and specificity were 82.9% and 78.0% during the prenatal period and 92.3% and 74.9% during the postpartum period, respectively. Figure 5 shows the corresponding ROC curves, and Table 4 provides the sensitivities and specificities of the paternal EPDS at various cut-off scores for detecting paternal perinatal depression. The scale also demonstrated a significant discriminatory ability, with AUC values of 0.877 (95% CI: 0.816–0.938; n = 387) for the prenatal period and 0.906 (95% CI: 0.839–0.973; n = 412) for the postpartum period. The AUC values during the prenatal period indicated excellent discrimination (AUC = 0.90–1.00), while the postpartum AUC values represented good discrimination (AUC = 0.80–0.89) (Fischer et al., 2003). These results support the utility of the scale for identifying depressive symptoms during both periods. Figure 6 presents scatter plots illustrating the relationship between the total paternal EPDS and CES-D scores for the prenatal and postpartum periods. Spearman’s rank correlation revealed a moderate and significant association during the prenatal period (ρ = 0.540, p <0.001) and a weaker but still significant association during the postpartum period (ρ = 0.538, p <0.001). The correlation results are presented in chronological order.

**Figure 5.**
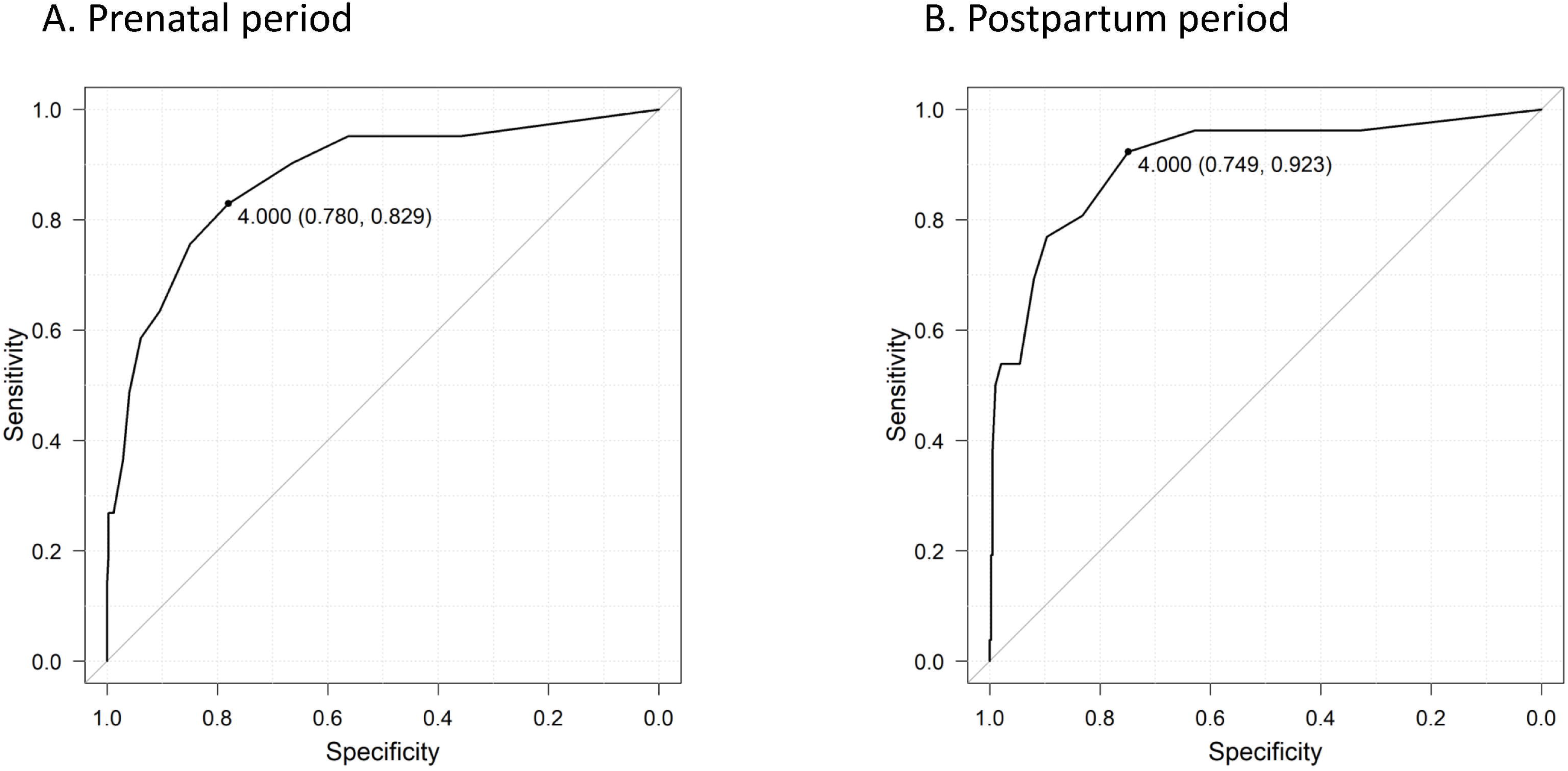
ROC curves of the paternal EPDS for detecting paternal perinatal depression (CES-D ≥16) during the prenatal and postpartum periods. The curves illustrate the diagnostic performance of the paternal EPDS for (A) the prenatal period (AUC = 0.877; 95% CI, 0.816–0.938) and (B) the postpartum period (AUC = 0.906; 95% CI, 0.839–0.973). The marked points indicate the optimal cut-off scores determined by the Youden Index, with coordinates presented as (Specificity, Sensitivity). The diagonal grey line represents the no-discrimination line. Abbreviations: AUC; Area under the curve; CI: confidence interval; CES-D: Center for Epidemiologic Studies Depression Scale; EPDS: Edinburgh Postnatal Depression Scale; ROC: Receiver operating characteristic.

**Figure 6.**
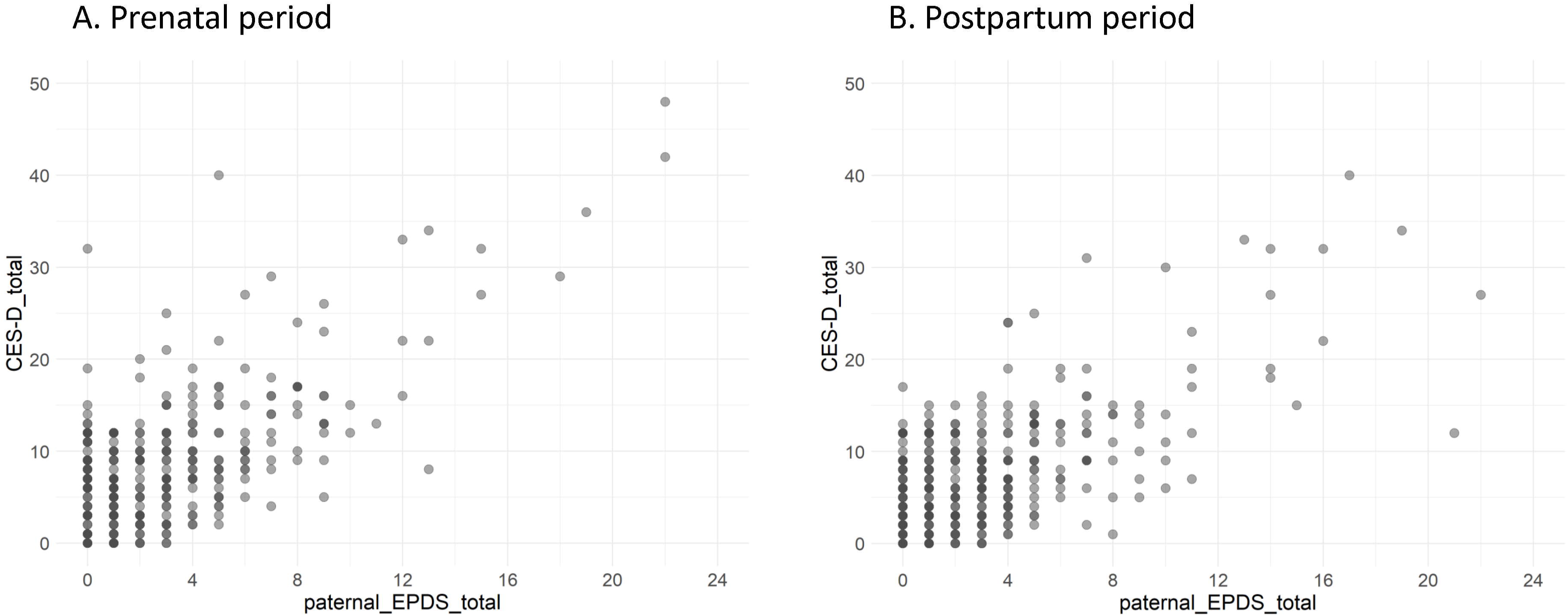
Scatter plots of the paternal EPDS scores versus paternal CES-D scores during the prenatal and postpartum periods illustrating the relationship between the paternal EPDS (completed by fathers) and CES-D (completed by fathers) for the (A) prenatal and (B) the postpartum periods. Each point represents an individual father. Spearman’s rank correlation revealed a moderate, significant association during both the prenatal period (ρ = 0.540, p < 0.001) and the postpartum period (ρ = 0.538, p < 0.001). Abbreviations: CES-D: Center for Epidemiologic Studies Depression Scale; EPDS: Edinburgh Postnatal Depression Scale.

**Table 4.** Sensitivity and specificity of the paternal EPDS at various cutoff scores for detecting paternal perinatal depression (CES-D ≥16) during the prenatal and postpartum period.

| paternal EPDS<br>Cutoff Score | Prenatal period |  | Postpartum period |  |
| --- | --- | --- | --- | --- |
|  | Sensitivity | Specificity | Sensitivity | Specificity |
| $\geq 0$ | 100.0% | 0.0% | 100.0% | 0.0% |
| $\geq 1$ | 95.1% | 36.1% | 96.2% | 33.2% |
| $\geq 2$ | 95.1% | 56.4% | 96.2% | 51.6% |
| $\geq 3$ | 90.2% | 66.5% | 96.2% | 63.0% |
| $\geq 4$ | 82.9% | 78.0% | 92.3% | 74.9% |
| $\geq 5$ | 75.6% | 85.0% | 80.8% | 83.2% |
| $\geq 6$ | 63.4% | 90.5% | 76.9% | 89.6% |
| $\geq 7$ | 58.5% | 93.9% | 69.2% | 92.0% |
| $\geq 8$ | 48.8% | 96.0% | 53.8% | 94.6% |
| $\geq 9$ | 36.6% | 97.1% | 53.8% | 96.4% |
| $\geq 10$ | 26.8% | 98.8% | 53.8% | 97.9% |
| $\geq 11$ | 26.8% | 99.4% | 50.0% | 99.0% |
| $\geq 12$ | 26.8% | 99.7% | – | – |
| $\geq 13$ | 19.5% | 99.7% | 38.5% | 99.5% |
| $\geq 14$ | – | – | 34.6% | 99.5% |
| $\geq 15$ | 14.6% | 100.0% | 19.2% | 99.5% |
| $\geq 16$ | – | – | 19.2% | 99.7% |
| $\geq 17$ | – | – | 11.5% | 99.7% |
| $\geq 18$ | 9.8% | 100.0% | – | – |
| $\geq 19$ | 7.3% | 100.0% | 7.7% | 99.7% |
| $\geq 21$ | – | – | 3.8% | 99.7% |
| $\geq 22$ | 4.9% | 100.0% | 3.8% | 100.0% |

“–” Values not calculable due to the absence of participants with paternal EPDS scores at those thresholds during the prenatal period.
The optimal cutoff score of 4 for both time points, as determined by the Youden index.
Abbreviations: EPDS: Edinburgh Postnatal Depression Scale; CES-D: Center for Epidemiologic Studies Depression Scale

## Discussion

This is the first study to validate the EPDS-P across the prenatal and postpartum periods while also demonstrating its diagnostic utility independent of maternal psychological and demographic factors. Logistic regression analyses revealed that maternal and paternal age, parity, gestational weeks or postpartum days, and the maternal EPDS and MIBS scores did not influence the EPSD-P scores, indicating that the EPDS-P remains a robust and valid screening tool, even when based on indirect observation. A key strength of this study lies in its relatively high response rates from a sample of the general population (71.3% [385/540] during pregnancy and 71.1% [411/578] during the postpartum period), suggesting a strong representativeness of the local perinatal population and reducing the risk of selection bias. Furthermore, the prevalence of depression did not differ between the sexes at any time: during pregnancy, 10.6% for men and 7.0% for women; postpartum, 6.1% for men and 5.4% for women (chi-square test, p >0.05). Among the 267 couples who responded at both time points, 78.6% (11/14) of the men who screened positive for postpartum depression already exhibited depressive symptoms during pregnancy. This finding underscores the importance of early screening and suggests that paternal depression often begins before childbirth and persists without timely intervention. The ROC analysis indicated fair discriminatory accuracy for the EPDS-P, with AUC values of 0.783 (95% CI: 0.700–0.866) during pregnancy and 0.746 (95% CI: 0.629–0.863) postpartum (Fischer et al., 2003). The optimal cut-off points were 3 during pregnancy and 4 during the postpartum period; the sensitivity and specificity values were 70.7% and 75.9% during pregnancy and 61.5% and 86.8% postpartum, respectively. The high specificity observed postpartum is particularly important for reducing false positives and improving resource allocation efficiency. However, moderate sensitivity at both time points raises concerns about potential under-detection; clinicians should still consider further assessments in at-risk fathers, even if their EPDS-P score is below the threshold. These AUC values are consistent with the findings of Fisher et al. (Fisher et al., 2012), who originally developed the EPDS-P, and support its applicability across cultural contexts. The fact that a similar diagnostic performance was observed in a Japanese community-based sample indicates the potential cross-cultural validity of this tool. Compared with the commonly used EPDS cut-off of 9 for women in Japan, the lower optimal thresholds for the EPDS-P in this study may reflect the inherent limitations of indirect screening. Research suggests that men often express depression through gendered symptoms, such as anger, irritability, overcommitment to work, substance use, or social withdrawal (Oliffe and Phillips, 2008; Oliffe et al., 2019), which may not be readily recognized by their female partners. This discrepancy highlights the potential misalignment between male-specific expressions of distress and female perception, highlighting the importance of incorporating sex-sensitive considerations into screening and intervention strategies (Paulson and Bazemore, 2010). Measures of masculine depression have been developed (Magovcevic and Addis, 2008), but there is limited research on this construct for perinatal fathers. Inclusion of “masculine depression” symptoms has been found to eliminate the sex disparities in the prevalence of depression between men and women in the general population(Martin et al., 2013), which may be relevant to the perinatal period. This calls for the development of a short-form screening measure of masculine depression for perinatal fathers.

However, it is noteworthy that the paternal EPDS, a self-reported measure, showed optimal cut-off values similar to those observed for the EPDS-P. This finding suggests that the overall maternal assessments were relatively accurate, indicating that female partners may be attuned to depressive symptoms in their male partners to a greater extent than was previously assumed. Another interpretation is that men may be less likely to overtly express psychological distress because of gender norms and social expectations, leading to similar underreported scores in both self- and indirect reports. Collectively, these findings support the practical utility of the EPDS-P in community-based settings, especially in Japan, where public health nurses routinely conduct perinatal home visits. Incorporating the EPDS-P into these services could allow for scalable, indirect screening of paternal mental health in a way that complements mother-centered care. While awareness of paternal perinatal mental health is increasing, health services and support systems remain heavily mother-centered. Preconception care has traditionally focused on optimizing women’s health (O’Connor et al., 2019), but growing evidence, including the findings of this study, supports the integration of paternal mental health screening into routine care. Doing so could offer benefits such as the early detection of at-risk fathers, psychoeducation on emotional transitions and coping strategies, and improved communication between partners, all of which may contribute to healthier family dynamics.

From a public health standpoint, paternal depression has been associated with increased couple conflict, lower parenting confidence, and difficulty in bonding with infants (Nishigori et al., 2020). Therefore, the failure to address men’s mental health needs may have long-term negative consequences for the entire family. The perinatal period represents a crucial opportunity for preventative interventions. An interesting finding in our study was the higher prevalence of paternal depressive symptoms during pregnancy compared to the postpartum period, which contrasts with previous research (Tokumitsu et al., 2020). One possible explanation is that study participation, including exposure to mental health information during the prenatal period, increased fathers’ self-awareness and willingness to seek help. This may have contributed to the reduction in postpartum symptoms. The observed decline in CES-D scores between pregnancy and postpartum supports the value of early engagement and education for expectant fathers (Chen et al., 2024; O’Connor et al., 2023). Although screening was offered and follow-up support was available, some at-risk men did not engage in the services. This gap may be due to structural barriers, such as work-related constraints, economic pressures, or stigma surrounding emotional vulnerability in men (Reay et al., 2023). Effective interventions must, therefore, go beyond screening to also address social stigma, increase service accessibility, and provide culturally appropriate support options (Reay et al., 2023; Wynter et al., 2024).

This study had several limitations. First, the study was conducted in a single regional municipality (Towada City, Aomori Prefecture), which may limit its generalizability to urban or socioeconomically diverse populations. Second, the EPDS-P is a screening tool, not a diagnostic tool, and it cannot differentiate between transient mood changes and clinical depression. Future studies incorporating structured clinical interviews could enhance the diagnostic validity. Third, the use of self-reported and partner-reported questionnaires increases the potential for reporting bias (Althubaiti, 2016). Finally, the partially longitudinal design limits causal inference. Comprehensive prospective studies that track fathers from preconception to the postpartum period are needed. Despite these limitations, this study provides important empirical evidence supporting the use of maternal indirect reporting in the Japanese perinatal mental health system. Continued validation and refinement, including digital adaptation and population-based deployment, are essential for maximizing the tool’s reach and impact. In summary, this study demonstrated the feasibility of indirect maternal assessments to screen for paternal depression during the perinatal period. The EPDS-P shows promise as a practical and valid tool, particularly when the direct participation of men is limited. Further validation, refinement, and integration into digital platforms are essential for advancing paternal mental healthcare in Japan.

## Conclusions

This study demonstrated that maternal assessments using the EPDS-P are a valid method for screening paternal perinatal depression during pregnancy and the postpartum period. The EPDS-P showed fair diagnostic accuracy and was not significantly influenced by maternal psychological characteristics, such as depressive symptoms or bonding scores. Additionally, paternal EPDS, completed directly by fathers, yielded similar cut-off values, suggesting that indirect maternal evaluations may reflect paternal mental health more accurately than previously assumed. These findings support the utility of the EPDS-P as an alternative screening tool, particularly in contexts where the direct assessment of fathers is not feasible. Integrating such validated indirect tools into routine care is a critical step toward establishing truly family-centered support systems, ultimately improving the mental and relational health of parents and promoting optimal child development globally.

## Declaration of Interest

### Data availability statement

Since our research dataset contained potentially sensitive information, ethical restrictions were applied. All requests for data should be directed to the Institutional Review Board of the Ethics Committee. Upon request, the board will decide whether to share the data. Contact information: Institutional Review Board, Ethics Committee, Dokkyo Medical University, 880 Kitakobayashi, Shimotsuga-gun, Mibu, Tochigi, 321-0293, Japan; Phone: +81-282-86-1111.

### Ethical approval

This study was conducted following the Declaration of Helsinki and the Ethical Guidelines for Medical and Health Research Involving Human Subjects in Japan. The Ethical Review Board of the Dokkyo Medical University reviewed and approved the research protocol on September 3, 2021 (Approval No. 2021-015).

### Informed consent

This prospective, longitudinal, observational study involved couples residing in Towada, Japan. Participants were recruited during perinatal home visits by public health nurses and midwives affiliated with the Towada City Community Health Center in the Aomori Prefecture. All participants received a full explanation of the study and provided written informed consent before participation.

## Acknowledgements

The authors would like to thank Professor Emeritus Tadaharu Okano and all the staff at the Child and Family Support Center in Towada City for their valuable contributions to this study.

## CRediT authorship contribution statement

Conceptualization: KT; Data curation: KT and TK; Formal analysis: KT; Funding acquisition: KT; Methodology: KT; Supervision: NS and NYF; Writing–original draft: KT; Writing–review and editing: NS, SDF, JT, KY and NYF. All authors approved the final version of the manuscript.

---

Figure S1. The Japanese version of the Edinburgh Postnatal Depression Scale-Partner assessment tool used in this study for the maternal rating of paternal depressive symptoms. The original scale is translated into Japanese with permission and collaboration from the original author, Dr. Sheehan David Fisher.

The Japanese version of the EPDS-Partner (EPDS-P) questionnaire, which was originally included as Figure S1, has been removed in accordance with medRxiv’s policy regarding non-English content. The original Japanese questionnaire will be available as a supplementary file in the peer-reviewed journal submission. For a copy, please contact the corresponding author.

